# Determinants of Time to Convalescence among COVID-19 Patients at Millennium COVID-19 Care Center in Ethiopia: A prospective cohort study

**DOI:** 10.1101/2020.10.07.20208413

**Authors:** Tigist W. Leulseged, Ishmael S. Hassen, Endalkachew H. Maru, Wuletaw C. Zewde, Nigat W. Chamesew, Kalkidan T. Yegile, Abdi B. Bayisa, Tariku B. Jagema, Teketel T. Admasu, Mesay G. Edo, Eyosias K. Gurara, Meseret D. Hassen, Etsegenet Y. Menyelshewa, Firaol M. Abdi, Mahlet B. Tefera, Siham S. Ali

**Author notes:** Corresponding author: Tigist W. Leulseged, Research development office, Millennium COVID-19 Care Center, Addis Ababa, Ethiopia, Department of Internal Medicine, St. Paul’s Hospital Millennium Medical College, Addis Ababa, Ethiopia.

## Abstract

**Aim:** To estimate time to recovery/convalescence and identify determinants among COVID-19 infected patients admitted to Millennium COVID-19 Care Center in Addis Ababa, Ethiopia.

**Methods:** A prospective cohort study was conducted among a randomly selected sample of 360 COVID-19 patients who were on follow up from 2^nd^ June to 5^th^ July 2020. Kaplan Meier plots, median survival times, and Log-rank test were used to describe the data and compare survival distribution between groups. Association between time to recovery/ convalescence and determinants was assessed using the Cox proportional hazard survival model, where hazard ratio, P-value, and 95% CI for hazard ratio were used for testing significance.

**Results:** The mean age of the participants was 32.4 years (± 12.5 years). On admission, 86.9 % had mild COVID-19, 78.6% were asymptomatic and 11.4% of the patients had a history of pre-existing co-morbid illness. The Median time to recovery/ convalescence among the study population was 16 days. The log-rank test shows that having non-mild (moderate and severe) disease, having one or more symptoms at presentation, and presenting with respiratory and constitutional symptoms seems to extend the time needed to achieve recovery. The Final Cox regression result shows that the presence of symptom at presentation was found to be a significant factor that affects time to recovery/ convalescence, the rate of achieving recovery/ convalescence among symptomatic patients was 44% lower than patients who were asymptomatic at presentation (HR= 0.560, 95% CI= 0.322-0.975, p-value=0.040).

**Conclusions:** Presence of symptom was found to be associated with delayed viral clearance. This implies symptomatic patients are more likely to be infectious because of the prolonged viral shedding in addition to the presence of a more concentrated virus in the upper respiratory tract that enhances the transmission. Therefore, attention should be given in the isolation and treatment practice of COVID-19 patients with regard to presence of symptom.

**Key Messages:**

- The study assessed the time to RT-PCR proven recovery (two consecutive negative viral shedding) and identified determinants that affected the time.
- Symptomatic infection is associated with delayed viral clearance.
- The finding of the study could be used to guide the isolation and treatment practice.

## INTRODUCTION

Currently, the world is facing a global pandemic with a new coronavirus SARS-CoV-2 (Severe Acute Respiratory Syndrome Corona Virus Type 2) causing an infectious disease named COVID-19 (Corona Virus Infectious Disease 2019). The infection primarily targets the human respiratory system and is mainly transmitted by respiratory droplets and close contact with an infected person ^1^.

As per the WHO daily situation report, the total cases of COVID-19 are increasing worldwide including the African region which got hit by the pandemic a little later. As of August 5, 2020, globally there were 18,354,342 confirmed cases and 696,147 deaths because of COVID-19 infection ^2^.

In Ethiopia since the first case of COVID-19 confirmed on March 13, 2020, there were a total of 19,875 confirmed cases and 343 deaths as of August 5, 2020. Most of the cases are from Addis Ababa, the capital of Ethiopia. At the beginning, most of them were imported cases from overseas travelers. Eventually, the transmission dynamics shifted to mainly contact with a confirmed case and then to cases with no identifiable contact or travel history. As per WHO daily situation report, currently the transmission is classified as community transmission ^2^.

Initially, little was known about the disease. As research continues, knowledge about the clinical, epidemiologic, laboratory, and radiologic characteristics of the disease is building up. The presenting symptoms and the severity of the disease seem to vary from place to place as it varies from individual to individual based on sex, age, and other characteristics reflecting the contribution of background characteristics of patients to the clinical presentation, severity, and outcome of the disease.

Regarding the spectrum of presenting symptoms, patients may not present with any symptom during the entire course of the disease while being infected (asymptomatic cases) or patients might present with respiratory and constitutional symptoms which are reported to be the commonest presentation in most setups. In addition, atypical presentations like hyposmia, hypogeusia, nasal congestion, rhinorrhea, sputum, cerebrovascular accidents, abdominal pain, vomiting, and diarrhea are also reported. These atypical symptoms are reported to be observed in the majority of patients in a study conducted in Europe but only in few patients in the United states making it difficult to generalize these symptoms as atypical for all setups ^3-9^.

The outcome of the disease varies based on individual underlying characteristics and health conditions and could be range from uneventful recovery to death^10-12^.

Disease severity and death are found to be affected by different factors including older age, presence of pre-existing comorbidities, fracture, lack of early improvement in arterial partial pressure of oxygen (PaO2) to fraction of inspired oxygen (FiO2) ratio, and abnormal prominent laboratory markers ^11,13-20^. The role of sex in the severity of the disease or outcome is not clear yet, one study point that there seems to be no difference in epidemiological characteristics with regard to sex, and in another study, it is reported that sex might have impact on the clinical presentation and prognosis showing worse prognosis among males ^21,22^.

Some studies are conducted around the world that investigates the duration of viral shedding. It is reported that the median duration from diagnosis up to negative viral shedding varies from place to place and could range from 14 to 48 days and the major determinants identified were age, sex, temperature at admission, time from symptom onset to admission, hospital length of stay, having symptomatic infection, fever, chest tightness, pneumonia, invasive mechanical ventilation and lymphocyte count less than 2.0 × 10^9^/L ^23-27^.

To tackle the COVID-19 threat in Ethiopia hospitals and health centers have been equipped with the capacity to treat COVID-19 patients and to isolate contacts with confirmed cases. In addition to this, to accommodate to the growing threat, a 1000 bed Millennium COVID-19 Care Center (MCCC) was organized by a makeshift of a previous Millennium hall/ Addis park.

In Ethiopia, as in most African countries, the number of incident cases, severity of the disease, and fatality rate is lower than the rest of the world. But the pattern might not continue like this considering the vulnerable nature of the society in the face of the triple burden of diseases; communicable diseases, non-communicable diseases, and injuries, and challenged health care system. Several studies have been conducted around the world, almost all outside Africa, to understand the disease pattern and outcome, and evidence point that there is a variation from place to place showing that there is still very little well-established knowledge about the disease. Also, the underlying population characteristics, economic status, health care structure, and endemic disease pattern of our country, and Africa in general, is different from other regions of the world making it difficult to predict and generalize findings based on the health care set up of other countries.

That said, understanding the disease course in terms of viral shedding time will assist in providing a tailored isolation and treatment practice.

Therefore, the objective of this study was to estimate time to recovery/convalescence and identify determinants among COVID-19 infected patients admitted at Millennium COVID-19 Care Center in Addis Ababa, Ethiopia.

## METHODS AND MATERIALS

### Study setting and period

The study was conducted to Millennium COVID-19 Care Center (MCCC), a makeshift hospital in Addis Ababa, the capital city of Ethiopia. The center is remodeled from the previous Millennium hall/ Addis park which was a multipurpose recreational, meeting, and exhibition center. The center has 1000 beds including an ICU with 10 mechanical ventilators.

### Study Design

The study design was institution based prospective cohort design. The follow up was made from 2^nd^ June to 5^th^ July, 2020.

### Source and Study Population

The source population was all patients admitted to MCCC with a confirmed diagnosis of COVID-19 using RT-PCR from referral centers and who were on follow up from 2^nd^ June to 5^th^ July. During this interval a total of 768 COVID-19 patients were seen at the Center.

The study population was all selected COVID-19 patients who were on treatment and follow up at MCCC from 2^nd^ June to 5^th^ July, 2020 and who full fill the inclusion criteria.

### Sample Size Determination and Sampling Technique

The sample size was determined by using sample size calculation formula of the survival method by considering the following statistical assumptions: 95% Confidence Interval (CI), power of 90%, survival probability of 0.5, 5% marginal error, 10% non-response rate and with finite population correction. The final sample size estimated for this study was 366.

To select the study participants, simple random sampling method using table of random numbers was employed.

### Inclusion and Exclusion Criteria

All selected COVID-19 patients who were on treatment and follow up at the MCCC from 2^nd^ June to 5^th^ July, 2020 were included.

### Operational Definitions

#### COVID-19 patient

any patient who tested positive for COVID-19 antigen as reported by a laboratory given mandate to test such patients by the Ethiopian Federal Ministry of Health ^28^.

#### Asymptomatic patient

any patient who tested positive for COVID-19 but does not have any symptoms. These patients are detected after isolation and contact tracing as done by the Ethiopian Public Health Institute (EPHI) ^28^.

#### COVID-19 severity score

was determined based on the WHO classification as follows ^29^.

- **Mild Disease:** Characterized by fever, malaise, cough, upper respiratory symptoms, and/or less common features of COVID-19 (headache, loss of taste or smell etc.)
- **Moderate Disease:** Patients with lower respiratory symptom/s. They may have infiltrates on chest X-ray. These patients are able to maintain oxygenation on room air.
- **Severe Disease:** These patients have developed complications. The following features can define severe illness.
  - Hypoxia: SPO2 ≤ 93% on atmospheric air or PaO2:FiO2 < 300mmHg (SF ratio < 315)
  - Tachypnea: in respiratory distress or RR>30 breaths/minutes
  - More than 50% involvement seen on chest imaging

#### Recovery/ convalescence

Recovery/ convalescence from Covid-19 infection as evidenced by two negative RT-PCR test done at least 24 hours apart.

#### Event

Achieving recovery/ convalescence from Covid-19 infection.

#### Censoring

Represents patients lost to follow-up, transferred out, died or completed the follow-up period without achieving the event.

#### Time to event or censoring

Time between date of laboratory sample taken which confirmed Covid-19 infection to recovery/ convalescence from Covid-19 infection or censoring (in days).

### Patient and public involvement

Patients or the public were not involved in the design, or conduct, or reporting, or dissemination plans of our research.

### Data Collection Procedures and Quality Assurance

An interviewer administered questionnaire that consists of the variables of interest was developed from the patient registration and follow up form and used to collect the necessary data from the patients and their medical charts.

The data collection tool was pretested on 5% of randomly selected patients and their medical charts and necessary amendment on the data collection tool was made.

Training on the basics of the questionnaire and data collection tool was given for ten data collectors (BSc nurses and General practitioners) and two supervisors (General practitioner and public health specialist) for one day.

Data consistency and completeness was checked before an attempt was made to enter the code and analyze the data.

### Data Management and Data Analysis

The collected data was coded and entered into Epi-Info version 7.2.1.0, cleaned and stored and exported into SPSS version 23 for analysis. Frequency tables, Kaplan Meier (KM) plots and median survival times were used to describe the data. Survival experience of different groups was compared using KM survival curves andLog-rank tests. Association between time to recovery/ convalescence and determinants was assessed using the Cox proportional hazard survival model, where hazard ratio, P-value and 95% CI for hazard ratio were used for testing significance and interpretation of results.

Univariate analysis was performed to calculate an unadjusted hazard ratio (UHR) and to screen out potentially significant independent variables at 25% level of significance. Association between the most relevant independent variables and the time to recovery/ convalescence from COVID-19 infection was assessedusing multivariable Cox proportional hazard survival model. Adjusted HR, P-value and 95% CI for HR were used to test significance and interpretation of results. Variables with p-value ≤ 0.05 were considered as statistically associated with time to recovery/ convalescence from COVID-19 infection in days. The basic assumptions of Cox Proportional Hazard model was tested using log- minus- log function.

## RESULT

### Censoring status and median time to recovery

Information was obtained from 360 COVID-19 patients among the 366 samples selected, making the response rate 98.4%.

Among the 360 patients, 132 (36.7%) of the patients achieved recovery/ convalescence while 228 (63.3%) were censored.

The median time to recovery/ convalescence was 16 days and it ranges from 7 to 21 days.

### Socio–demographic and pregnancy related variables and censoring status

The mean age of the participants was 32.4 years (± 12.5 years). The minimum and the maximum age of the patients were 1 and 75 years respectively. Majority of the patients (55.8 %) were males and the rest (44.2 %) were females and 337 of the patients (93.6 %) were from Addis Ababa. Only 11 (3.1%) of the patients were health care professionals. There were three pregnant women, two at first trimester and one at third trimester.

The proportion of patients who achieved recovery among males (39.8%) was higher than females (32.7%). Higher proportion of patients from outside Addis Ababa (52.2%) achieved recovery compared with patients from Addis Ababa (35.6%). Majority of the health care professionals (90.9%) and all the three pregnant were censored. (**Table 1**)

**Table 1:** Baseline socio-demographic, behavioral, medical and disease related variables and censoring status among COVID-19 patients (n=360)

| Variable | Censoring status |  | Total (%) |
| --- | --- | --- | --- |
|  | No of censored (%) | No of event (%) |  |
| **Sex** |  |  |  |
| Female | 107 (67.3) | 52 (32.7) | 159 (44.2) |
| Male | 121 (60.2) | 80 (39.8) | 201(55.8) |
| **Place of residence** |  |  |  |
| Outside Addis Ababa | 11 (47.8) | 12 (52.2) | 23 (6.4) |
| Addis Ababa | 217 (64.4) | 120 (35.6) | 337 (93.6) |
| **Health care professionals** |  |  |  |
| No | 218 (62.5) | 131(37.5) | 349 (96.9) |
| Yes | 10 (90.9) | 1 (9.1) | 11 (3.1) |
| **Pregnancy status** |  |  |  |
| No | 32 (20.5) | 124 (79.5) | 156 (99.2) |
| Yes | 3 (100) | 0 | 3 (0.8) |
| **How patient contracted the disease** |  |  |  |
| Non community transmission * | 74 (70.5) | 31(29.5) | 105 (29.2) |
| Community transmission | 154 (60.4) | 101(39.6) | 255 (70.8) |
| **Pre-existing co-morbid illness** |  |  |  |
| No | 198 (62.1) | 121(37.9) | 319 (88.6) |
| Yes | 30 (73.2) | 11(26.8) | 41 (11.4) |
| **Substance use\*** |  |  |  |
| No | 223 (62.8) | 132(37.2) | 355 (98.6) |
| Yes | 5 (100) | 0 | 5 (1.4) |
| **Drug use in 14 days (ACEIs, ARBs and NSAIDs)** |  |  |  |
| No | 222 (62.7) | 132 (37.3) | 354 (98.3) |
| Yes | 6 (100) | 0 | 6 (1.7) |
| **Presence of symptom/s** |  |  |  |
| No | 168 (59.4) | 115 (40.6) | 283 (78.6) |
| Yes | 60 (77.9) | 17 (22.1) | 77 (21.4) |
| **Respiratory symptoms\*** |  |  |  |
| No | 175 (59.5) | 119 (40.5) | 294 (81.7) |
| Yes | 53 (80.3) | 13 (19.7) | 66 (18.3) |
| **Constitutional symptoms\*** |  |  |  |
| No | 192 (61.1) | 122 (38.9) | 314 (87.2) |
| Yes | 36 (78.3) | 10 (21.7) | 46 (12.8) |
| **Neurologic symptoms\*** |  |  |  |
| No | 227 (63.6) | 130 (36.4) | 357 (99.2) |
| Yes | 1 (33.3) | 2 (66.7) | 3 (0.8) |
| **Gastro intestinal symptoms\*** |  |  |  |
| No | 224 (63.5) | 129 (36.5) | 353 (98.1) |
| Yes | 4 (57.1) | 3 (42.9) | 7 (1.9) |
| **COVID-19 Severity** |  |  |  |
| Mild | 191 (61) | 122 (39.0) | 313 (86.9) |
| Moderate | 34 (77.3) | 10 (22.7) | 44 (12.2) |
| Severe | 3 (100) | 0 | 3 (0.8) |

**Non community transmission\*:** includes contact with a diagnosed person, working in a center caring for COVID-19 patients, history of travel outside of Ethiopia and contact with a traveler
**Substance use\*:** includes cigarette smoking, shisha smoking and khat chewing.
**Respiratory symptoms\*:** includes dry cough (13.6%), cough with sputum production (5.6%), sore throat (6.1%), runny nose (3.1%), chest pain (3.9%) and shortness of breath (1.9%).
**Constitutional symptoms\*:** includes fever (6.4%), myalgia (2.8%), arthralgia (2.2%), malaise (4.7%) and headache (6.4%).
**Neurologic symptoms\*:** includes altered consciousness (0.3%) and seizure (0.6%).
**Gastrointestinal symptoms\*:** includes abdominal pain (0.6%), vomiting/ nausea (1.1%) and diarrhea (0.8%).

### Contact history, pre-existing history of Co-morbid illness, substance use and medication intake history related variables and censoring status

Close to two third of the patients (70.8%) acquired the disease through community transmission and the rest (29.2%) were through a known contact or travel history. Forty one (11.4%) of the patients had a history of pre-existing co-morbid illness. The commonest co-morbid illness history was hypertension (5%) followed by type 2 diabetes mellitus (2.2%), HIV (1.9%), bronchial asthma (1.1%), seizure disorder (0.6%), chronic liver disease (0.3%) and tuberculosis (0.3%). Five of the patients (1.4%) had a history of substance use including cigarette and shisha smoking and khat chewing. Regarding drug used within 14 days of admission, ACEIs, ARBs and NSAIDs use were reported by one (0.3%), one (0.3%) and five (1.4%) of the patients, respectively.

The proportion of patients who achieved recovery was higher among those who contracted the disease through community transmission (39.6%) and those with no pre-existing co-morbid illness (37.9%) compared with those who contracted the disease through non-community transmission (29.5%) and those with pre-existing co-morbid illness (26.8%), respectively. All the six patients who have a history of drug intake within 14 days of admission were censored. (**Table 1**)

### Presenting symptom and COVID-19 severity related variables and censoring status

More than two third (78.6%) of the patients were asymptomatic at presentation and the rest (21.4%) were symptomatic. The commonest presenting symptom was respiratory symptoms (18.3 %) followed by constitutional (12.8%), Gastrointestinal (1.9%) and neurologic (0.8%) symptoms. Majority of the patients (86.9 %) had mild COVID-19 severity score at admission and the rest had moderate (12.2%) and severe (0.8%) disease at presentation.

The proportion of patients who achieved recovery was higher among those with no presenting symptom at admission (40.6%) than those with one or more symptom at admission (22.1%).

The proportion of patients who achieved recovery was higher among those who did not have respiratory (40.5%) and constitutional (38.9%) symptoms compared with those who presented with respiratory (19.7%) and constitutional (21.7%) symptoms. On the other hand, higher proportion of patients with neurologic (66.7%) and gastrointestinal symptoms (42.9%) has achieved recovery compared with those with no neurologic (36.4%) and gastrointestinal (36.5%) symptoms at presentation. A greater proportion (39.0%) of patients with mild COVID-19 severity score has achieved recovery compared with the moderate cases (22.7%). All the three severe cases were censored. (**Table 1**)

### Comparison of survival experience

A log rank test was used to assess difference in the survival distribution among groups. Accordingly, the survival time was significantly different among the different groups in COVID-19 severity score, presence of symptom, respiratory and constitutional symptoms (p-values < 0.05). Having non mild (moderate and severe) disease and having one or more symptom at presentation seems to extend the time needed to achieve recovery. The time needed to achieve recovery was longer among patients with non mild disease (19.3 days) compared with those with mild disease (17.9 days) (X^2^_(1)_= 7.841, P-value= 0.005). The survival time showed that those patients who presented with one or more symptom achieved recovery in a relatively longer time (19.2 days) than those who had no symptom at presentation (17.8 days). As shown in **Figure 1** the KM survival function graph also showed that those with mild disease and those with no symptom at presentation had a favorable survival (time to achievement of recovery/ convalescence) experience. The right panel of the figures shows that, the instantaneous chance of achieving recovery increases for both COVID-19 severity groups and presenting symptom groups as the duration of treatment increases.

**Figure 1:**
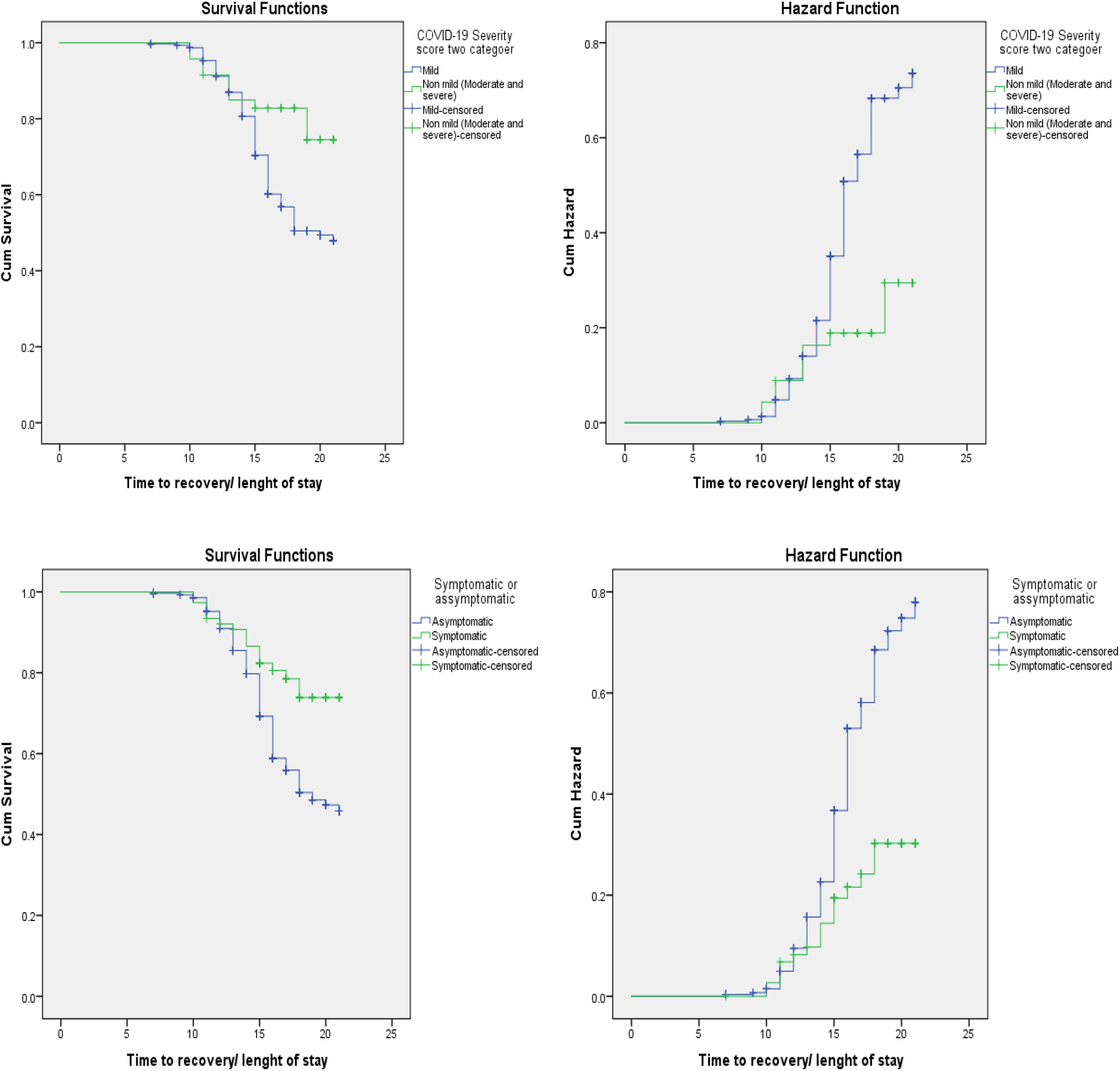
Survival and Hazard functions of COVID-19 severity score and presence of symptom by time

Similarly, the time needed to achieve recovery seemed longer among patients who presented with respiratory symptom (19.3 days) and constitutional symptom (19.4 days) compared with those who didn’t have respiratory symptom (17.9 days) and those who didn’t have constitutional symptom (17.9 days), respectively.

On the other hand, the survival time did not show statistically significant difference among the different groups classified by sex, place of residence, health care worker, how patient contracted the disease and pre-existing co-morbid illness (all p-values >0.05). (**Table 2)**

**Table 2:**
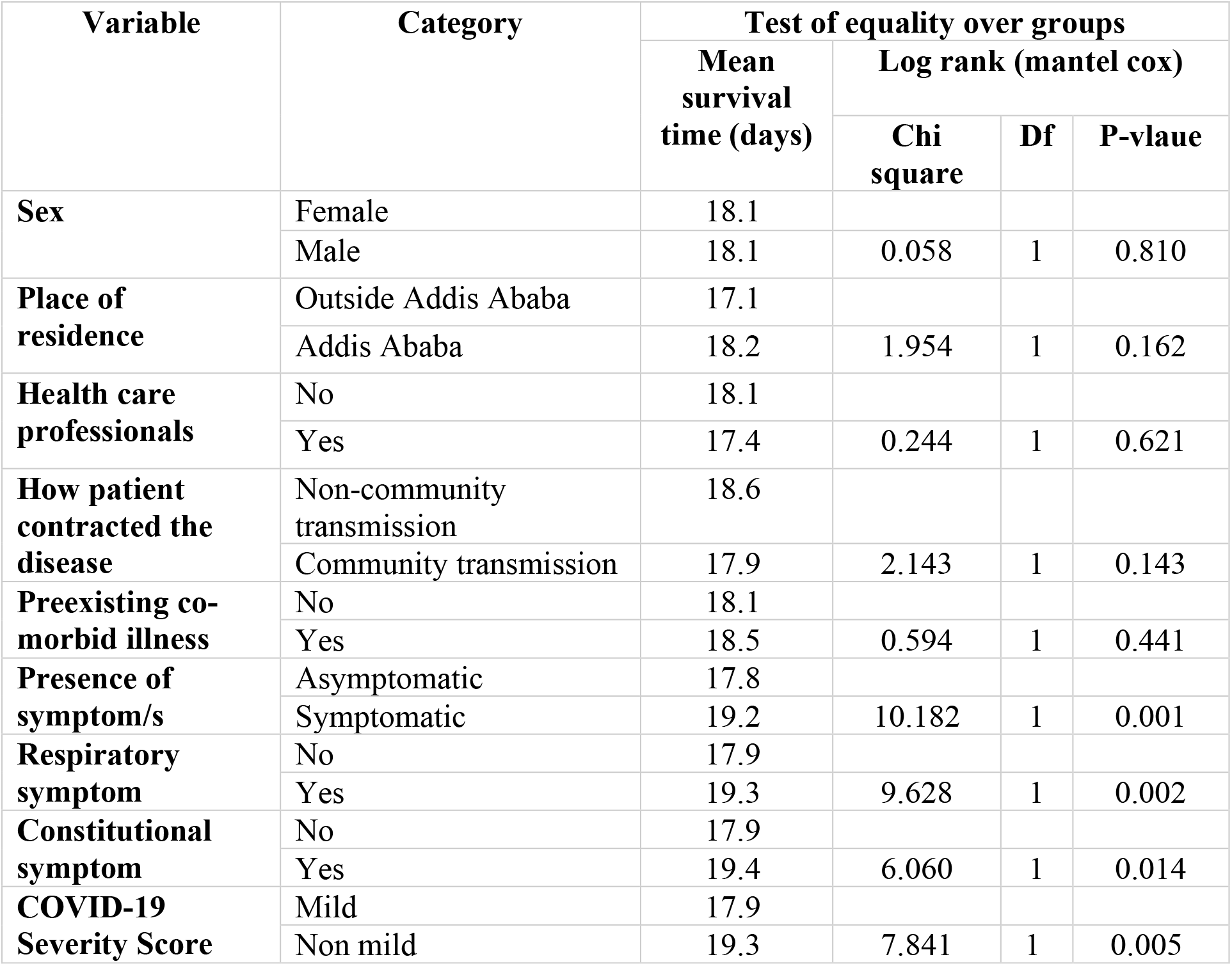
Comparison of recovery from COVID-19 among COVID-19 patients (n=360)

### Results of Multivariable Cox Proportional Hazard Model

The fundamental assumption of Cox Proportional Hazard model, which is proportional hazards assumption, was tested using Log-minus-Log function. Parallel lines between groups indicate proportionality ^30^. **Figures 2** reveals that the survival curves seem parallel throughout the study time. These plots show reasonable fit to the proportional hazard assumption.

**Figure 2:**
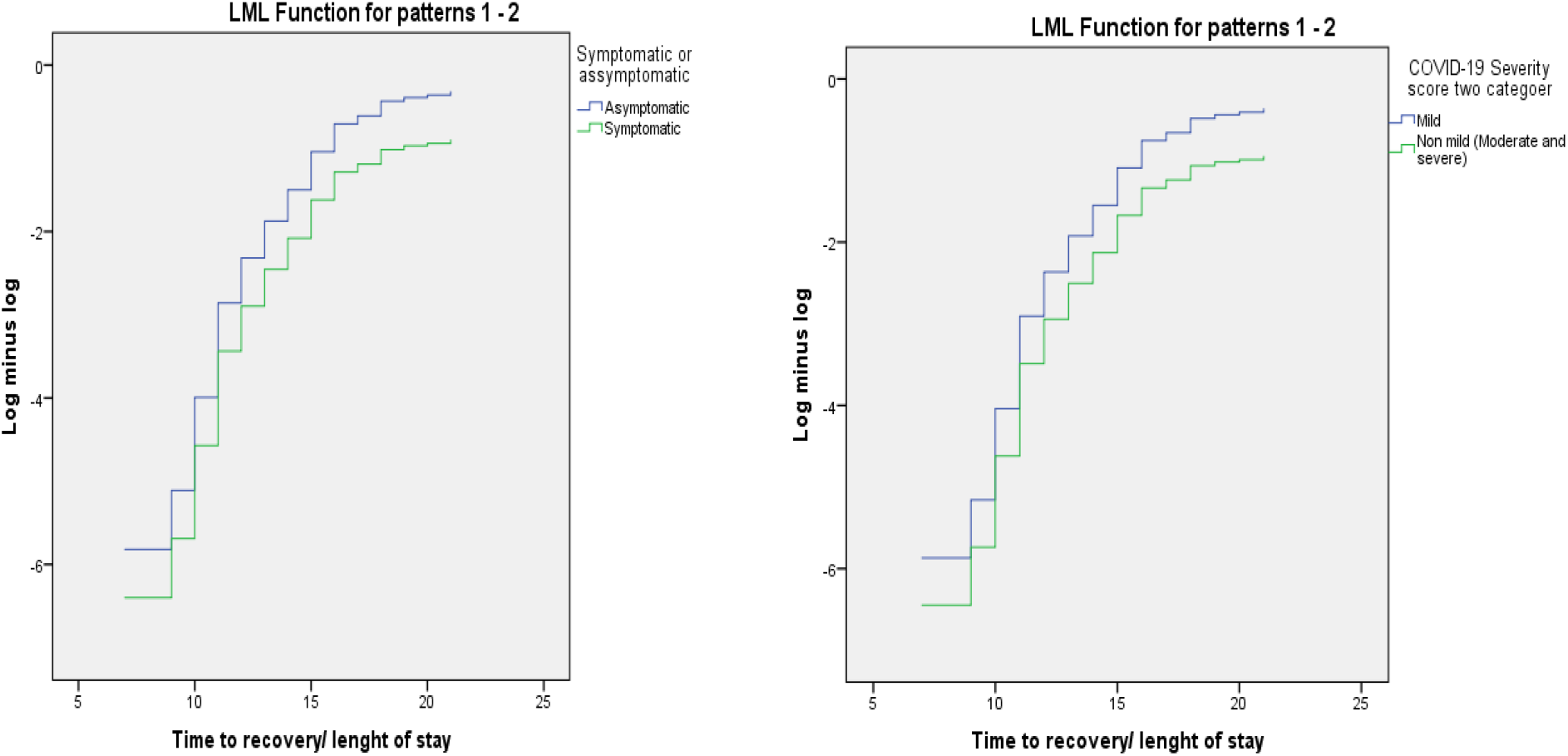
Log minus Log function for presence of symptom and COVID-19 severity score

Based on the result of the Univariate analysis at 25% level of significance and the significant variables identified from literatures, the following variables were included in the final regression model; age, sex, pre-existing co-morbid illness/s, presence of symptom and COVID-19 severity.

Presence of symptom at presentation was found to be significantly associated with time to recovery/ convalescence in the multivariable Cox proportional hazard model at 5% level of significance. Accordingly, after adjusting for other covariates, the rate of achieving recovery/ convalescence among symptomatic patients was 44% lower than patients who were asymptomatic at presentation (HR= 0.560, 95% CI= 0.322-0.975, p-value=0.040). This implies that the time needed to reach to recovery among symptomatic patients was significantly longer compared with asymptomatic patients. (**Table 3**)

**Table 3:** Results for the final Cox proportional hazard model among COVID-19 patients (n=360)

| Variables | CHR (95% CI) | AHR | 95.0% CI for AHR | P-value |
| --- | --- | --- | --- | --- |
| **Age** | 0.999 (0.986, 1.012) | 1.002 | (0.988, 1.016) | 0.796 |
| **Sex** |  |  |  |  |
| Female |  | 1 |  |  |
| Male | 1.042 (0.735, 1.478) | 1.077 | (0.758, 1.532) | 0.678 |
| **Pre existing/ concomitant Co-morbid illness** |  |  |  |  |
| No |  | 1 |  |  |
| Yes | 0.762 (0.411, 1.413) | 0.852 | (0.449, 1.616) | 0.623 |
| **Presence of Symptom** |  |  |  |  |
| Asymptomatic |  | 1 |  |  |
| Symptomatic | 0.458 (0.275, 0.763) | 0.560 | **(0.322, 0.975)** | **0.040\*** |
| **COVID-19 Severity** |  |  |  |  |
| Mild |  | 1 |  |  |
| Non mild | 0.422 (0.221,0.805) | 0.560 | (0.272, 1.154) | 0.116 |

**Note:** CHR, Crude Hazard ratio; AHR, Adjusted Hazard ratio; CI, Confidence interval; \*Statistically significant

## DISCUSSION

In this study, we assessed the determinants of time to convalescence among COVID-19 patients who were admitted at MCCC. The log-rank test shows that having non-mild (moderate and severe) disease, having one or more symptom at presentation, and presenting with respiratory and constitutional symptoms seems to extend the time needed to achieve recovery. This implies that having a more severe disease category and developing symptomatic disease are associated with delayed biochemical recovery from the disease and extended infectiousness period which puts close contact at risk.

The median time to recovery/ convalescence was 16 days. There is no a set standard or target that is set to be used as a reference internationally or nationally. This finding is comparable with results from other studies that reported a median duration of 14, 15, and 17 days in different countries and different age groups^23-26^.

According to the Cox proportional hazard regression model, the presence of symptom at presentation was found to be a significant factor that determines the time to recovery/ convalescence.

The rate of achieving recovery/ convalescence among symptomatic patients was 44% lower than patients who were asymptomatic after being adjusted for other variables. This means symptomatic patients showed a relatively delayed viral clearance compared to asymptomatic patients. This could be because most of the symptomatic patients in the current study had respiratory tract symptoms that could have increased the likelihood of detecting the virus in the upper respiratory tract sample. This finding is also in accordance with a study conducted in Wuhan where having symptom was found to be a significant predictor of duration of viral shedding, with a prolonged viral shedding among patients who present with symptomatic infection^26^.

On the other hand, factors that are identified to be significant determinants of convalescence like age, sex, presence of co-morbid illness, and disease severity didn’t show any significant association with time to convalescence.

In this study, the majority of the participants (63.3%) were censored due to referral to other centers, and due to the relatively short study period. The upper limit of the observation period is chosen because of the change in the patient discharge criteria which focused on a clinical decision in addition to the RT-PCR result per se which made us unable to get two consecutive negative RT-PCR results for each patient. This might have reflected on the median time obtained. Therefore, the result has to be interpreted with this limitation in mind.

## CONCLUSION

The median time to recovery/ convalescence among COVID-19 patients admitted at Millennium COVID-19 Care Center was found to be 16 days. The presence of symptoms was found to be associated with delayed viral clearance. This implies that symptomatic patients are more likely to be infectious because of the prolonged viral shedding in addition to the presence of a more concentrated virus in the upper respiratory tract that enhances the transmission. Therefore, attention should be given in the isolation and treatment practice of COIVD-19 patients with regard to the presence of symptom. Furthermore, to guide the local practice with better evidence, further multicenter study with prolonged observation is recommended.

## Data Availability

All relevant data are available upon reasonable request.

## Declaration

### Ethics approval and consent to participate

The study was conducted after obtaining ethical clearance from St. Paul’s Hospital Millennium Medical College Institutional Review Board. Written informed consent was obtained from the participants. The study had no risk/negative consequence on those who participated in the study. Medical record numbers were used for data collection and personal identifiers were not used in the research report. Access to the collected information was limited to the principal investigator and confidentiality was maintained throughout the project.

### Consent for publication

Not applicable

### Competing interests

The authors declare that they have no known competing interests

### Funding source

This research did not receive any specific grant from funding agencies in the public, commercial, or not-for-profit sectors.

### Authors Contribution

TWL conceived and designed the study, revised data extraction sheet and drafted the initial manuscript. KTY, ABB and TBJ designed data extraction sheet. All authors contributed to the conception and obtained patient data. TWL, KTY and ABB performed statistical analysis. All authors undertook review and interpretation of the data. All authors revised the manuscript and approved the final version.

## Acknowledgment

The authors would like to thank St. Paul’s Hospital Millennium Medical College for facilitating the research work.

## Availability of data and materials

All relevant data are available upon reasonable request.

## Notes

### Competing Interest Statement

The authors have declared no competing interest.

### Author Declarations

The study was conducted after obtaining ethical clearance from St. Paul's Hospital Millennium Medical College Institutional Review Board.

